# Therapists’ perspectives on the use of an overground lower-limb exoskeleton in physiotherapy sessions for minimally ambulatory children with cerebral palsy: a qualitative study

**DOI:** 10.64898/2026.09.08.26361200

**Authors:** Ledycnarf Januario de Holanda, Stefanie S. Bradley, F. Virginia Wright

## Abstract

**Background and objective:** Children with cerebral palsy (CP) generally experience more sedentary lifestyles than neurotypical peers. Powered overground exoskeletons offer stable and assisted stepping that have been shown to lead to functional and exercise-linked participation opportunities. This study aimed to explore physiotherapy team experiences, perceived benefits and challenges when using an overground lower-limb exoskeleton in physiotherapy sessions with minimally ambulatory children with CP.

**Methods:** Qualitative descriptive study within a pre–post feasibility trial involving 10 children with CP (GMFCS Level IV; 4-6 years) who received lower limb exoskeleton-based physiotherapy twice weekly over six weeks in school or outpatient setting. Physiotherapist (PT; n=5) and physiotherapy assistant (PTA; n=4) intervention team dyads were interviewed following completion of the exoskeleton intervention with their assigned child. Semi-structured interviews were conducted over Zoom. Coding of interview transcripts was followed by inductive thematic analysis.

**Results:** There were 10 PT/PTA dyad interviews linked specifically to 10 children’s intervention experiences in the exoskeleton study. Three themes were developed: 1) exoskeleton-based sessions provided additional exercise/gross motor options and may be a valuable adjunct to conventional therapy; 2) exoskeleton-based sessions were enjoyable, supporting enhanced inclusion and autonomy; 3) exoskeleton-based physiotherapy requires new clinical skills, added resources and individualized goal-based thinking. Therapists described exoskeleton-based therapy as a complement to, rather than a replacement for, conventional physiotherapy, since it addresses only a subset of a child’s motor goals. Therapists identified staffing, training, environmental accessibility, and equitable resource allocation as important considerations for clinical implementation.

**Discussion:** This study provides PTs/PTAs’ perspectives on their first use of an overground exoskeleton across school and outpatient settings with children with CP. Findings highlight the perceived value and logistical considerations of lower-limb exoskeleton use within physiotherapy sessions with minimally ambulatory children with CP. The findings were mapped onto the seven facets of the Theoretical Framework of Acceptability for new interventions and may inform implementation planning. Transferability to other settings should be considered in light of differences in resources, funding structures, and organizational contexts. Future research exploring ethical considerations, service delivery models, and clinical protocols is needed to support responsible integration into therapy settings across diverse clinical contexts.

## INTRODUCTION

Children and adolescents (hereafter referred to as children) with cerebral palsy (CP) generally experience more sedentary lifestyles than neurotypical peers, participating less in self-care, physical activities, play, and leisure [1–3]. Benefits of standing and ambulation can include improved bone health, prevention of muscle contractures and obesity, enhanced cardiovascular function, greater autonomy, and additional opportunities to participate in physical activities with family and peers [4–7]. For children who primarily use wheelchairs for mobility, but are able to take steps (e.g., those in Gross Motor Function Classification System [GMFCS] Level IV [8]), manual supported-stepping walkers (e.g., Rifton Pacer and Mustang Gait Trainer) provide an option of trunk-supported, upright weight bearing mobility [6] and associated physical activity. However, functional use of these walkers is often severely limited by inefficient gait patterns [9], postural control challenges, reduced strength, and high energy cost [10,11].

Assistive gait devices in the form of overground powered exoskeletons (i.e., portable robotic systems that support ambulation over indoor/outdoor distances) have been designed to reduce these challenges [12]. Exoskeletons such as the CPWalker, Atlas 2030, Angel Legs M-20, Explorer, and Trexo [13–17] have been built specifically for minimally ambulatory children with neuromotor conditions with the aim to provide new functional and exercise-linked walking opportunities in home and community environments. These overground exoskeletons can be also used within gait-focused physiotherapy sessions, similar to what has been done over the past two decades with treadmill-based, tethered (stationary) exoskeletons such as the Lokomat [18]. These stationary devices provide high intensity, gait-based exercise, and there is evidence of positive impact on spatiotemporal aspects of gait in children with CP, however conclusions are inconsistent regarding extent and breadth of functional gait-related benefits [19–21].

While powered overground exoskeletons provide similar repetitive supported stepping and gait-based exercise to stationary exoskeletons, they also allow sensory environmental interaction and whole task participation opportunities that may confer added advantages [12]. However, effectiveness evidence is early stage (i.e., pilot/feasibility studies), with much of the published research thus far focused on the Trexo lower limb exoskeleton (Trexo Robotics, Mississauga, Canada) [17,22–26]. This maximum support overground device integrates powered orthotic legs within a modified Rifton Pacer. The Trexo was introduced first into home use context with a child’s caregiver operating its tablet-based robotic control system and guiding its steering via a walker-mounted handle. In addition to facilitating gait [17,23], home use pre-/post-test studies have suggested associated benefits in sleep quality, bowel function, head control and knee spasticity [17,22]. A 12-week Trexo home-based study demonstrated accomplishment of individualized goals and parent reported benefits in social experiences and family interactions, although challenges were noted with achieving weekly use targets [26].

When considering use of robotic technologies within clinical practice, successful integration requires attention to implementation factors including, but not limited to training, safety, usability, goal alignment, accessibility, and transferability among eligible clients [27]. As well, there are complex human-exoskeleton interaction aspects for physiotherapists (PTs) to navigate [28]. From an adoption perspective, PTs are often sceptical about operational, resource, and cost-associated demands of exoskeleton implementation [27,29]. It is thus essential, in tandem with effectiveness evaluations, to understand their viewpoints on device usefulness, suitability, and operational barriers to ensure comfortable technology use in clinical sessions and optimize impact within a clinically viable service delivery approach [30,31].

Thus, the goal of this qualitative study was to explore PT and physiotherapy assistants’ (PTA) perspectives regarding the use of a lower-limb exoskeleton within physiotherapy sessions. This project was embedded in a pre-post intervention study [32] that explored the feasibility and functional effects of physiotherapy-based overground exoskeleton use (the Trexo, as described above) with 10 minimally ambulatory children with CP (i.e., in GMFCS level IV [8]) at our pediatric rehabilitation centre. The underlying thinking was that exoskeleton use within this intervention context could provide new opportunities to work on individualized gross motor and functional gait activity goal areas. Each child in the study received exoskeleton-based physiotherapy sessions twice weekly over 6 weeks, delivered in our centre’s school setting (n=5 participants) or out-patient program (n=5). The focus and format of these sessions is summarized in Table 1.

**Table 1.** Session design for outpatient and school physiotherapy-based exoskeleton sessions in the overarching feasibility study [25].

|  | Exoskeleton Outpatient Therapy | Exoskeleton School Therapy |  |
| --- | --- | --- | --- |
|  | Individual Sessions | School gym class sessions | School Individual Sessions |
| Frequency | Twice per week | Once per week | Once per week |
| Duration | 50 minutes (30-40 minutes active treatment) | 40 minutes (20-30 minutes active treatment) | 40 minutes (20-30 minutes active treatment) |
| Team | Child’s PT/PTA team pre-session<br>Trexo setup done by research team | Child’s PT/PTA team, pre-session<br>exoskeleton setup done by research team | Child’s PT/PTA team, pre-session<br>exoskeleton setup done by research team |
| Environment | • Gait lab, hospital hallways, or outdoors during summer months | • School gymnasium<br>• Sometimes transitioning to classroom after gym class | • School therapy activity centre<br>• School hallways<br>• Outside the school (adjoining hospital hallways, outdoors during summer months) |
| Kind of activities planned | • Individual activities focused on gait quality, endurance, upper body control, upper limb use (motor learning approach taken) | • Activities planned by gym teacher for whole class and exoskeleton use was integrated into this<br>• Interaction and socialization with peers while doing activities in the exoskeleton | • Individual activities focused on gait quality, endurance, upper body control, upper limb use (motor learning approach taken) |
| Parent presence/involvement | • Parents present during all sessions, supporting communication, interpretation of fatigue and comfort, and allowing direct observation of the child’s participation | • Parents and siblings were not present at school sessions<br>• Parents accessed the intervention indirectly through regular progress communication with the PT, attendance at one exoskeleton school session, and review of video footage (1–2 sessions) via a confidential web-link | • Parents and siblings were not present at school sessions<br>• Parents accessed the intervention indirectly through regular progress communication with the PT, attendance at one exoskeleton school session, and review of video footage (1–2 sessions) via a confidential web-link |

Our feasibility study results [25] showed the exoskeleton-based therapy was safe, feasible within the research context, and associated with achieving the intervention-focused goals set for use in our school and outpatient session setting. The aims of this qualitative study, considered alongside these feasibility findings, were to further inform decisions on future clinical implementation of the exoskeleton at our facility, offer insights for other centres considering exoskeleton use, and serve as a resource for future hybrid effectiveness-implementation research involving this and similar pediatric exoskeletons. The study addressed the following research question: What are the physiotherapy team experiences, perceived benefits and challenges of overground lower-limb exoskeleton use during physiotherapy sessions with young, minimally ambulatory children with CP?

## MATERIALS AND METHODS

### Study design

This qualitative descriptive study [33,34] consisted of semi-structured interviews with the outpatient and school PT/PTAs who provided the exoskeleton-based therapy sessions in the feasibility study [25]. Each PT and PTA was invited by the study research assistant (RA) to an interview after completion of their study child’s exoskeleton intervention. Informed consent was obtained from the PT and PTA for each interview done. The project received ethics approval from the Holland Bloorview Kids Rehabilitation Hospital (REB#0523) and University of Toronto (REB# 00044118).

### Lower limb exoskeleton therapy

A medium-sized Trexo Plus device was used for physiotherapy sessions, facilitated by a two-person PT/PTA team that received device training prior to the study start. For all child exoskeleton users, the Trexo robotic legs and child were oriented outwards (facing out of the open side of the Rifton frame) to physically maximize participation during physiotherapy [25].

### Child exoskeleton users and session details from the overarching feasibility study

Table 2 summarizes key gross motor, mobility, and communication characteristics of the 10 children (GMFCS Level IV; aged 4-6 years) in the feasibility study [25]. Children were either between therapy blocks (out-patient participants) or had their usual physiotherapy replaced by these exoskeleton sessions (school participants).

**Table 2.** Characteristics of children enrolled in the overarching exoskeleton feasibility study [25].

| **Therapy Setting** | **Child Exoskeleton User (pseudonym)** | **Manual Stepping Device** | **Movement with Manual Stepping Device** | **GMFM-66 Score<sup>1</sup>** | **MACS<sup>2</sup>** | **CFCS<sup>3</sup>** |
| --- | --- | --- | --- | --- | --- | --- |
| Outpatient | Ryan | Rifton Pacer | Stepping up to 100m with maximum effort; some steering from PT | 34.9 | III | I |
| School | Joseph | Rifton Pacer | Minimal independent movement; requires maximal assistance from PT | 26 | IV | IV |
| School | Samia | Mustang Walker | 2-3 independent steps with maximum effort | 31.8 | IV | IV |
| School | Gabriela | Mustang Walker | Stepping up to 100m with maximum effort; steering from PT | 30.6 | III | II |
| School | Ana | Mustang Walker | Functional in manual device, fast pace with some steering from PT | 26 | V | II |
| Outpatient | Jasmine | No walker | No walker at time of study | 4.1 | V | V |
| Outpatient | Sophie | Rifton Pacer | 2-3 independent steps with maximum effort | 22.7 | IV | V |
| Outpatient | Adil | Rifton Pacer | Functional in manual device, fast pace with some steering from PT | 38.8 | III | III |
| Outpatient | Mark | Rifton Pacer | Stepping up to 100m with maximum effort; some steering from PT | 30.6 | III | V |
| School | Jaden | Rifton Pacer | Highly functional in manual device, slight steering help from PT | 50.9 | II | III |

Gross Motor Function Measure [8]; <sup>2</sup>Manual Ability Classification System [35]; <sup>3</sup>Communication Function Classification System [36].

Therapeutic activity focus of the motor-learning based exoskeleton sessions was individualized to children’s abilities and gross motor/functional goals. These goals originated from their most recent physiotherapy care and were refined into Canadian Occupational Performance Measure (COPM) goals [37] (Table 3a) by study assessors together with parents to align with possible outcomes associated with exoskeleton therapy [25]. Related new goals, many of which focused on exoskeleton or supported-stepping walker user abilities, were set by the child’s study PT using Goal Attainment Scaling [38] and evaluated post-intervention by this same PT (Table 3b). These goals are provided in this paper to give clear context to the PTs’/PTAs’ comments about goals of their exoskeleton session and observed outcomes specific to each child. Individualized therapy activities that were done in the exoskeleton sessions to work towards these goals included, but were not limited, to: walking distance (endurance), walking speed (fitness), gait quality, neck/trunk control, upper limb use, and dual tasking functional activities, as detailed in the feasibility study publication [25].

**Table 3a.** Examples of Canadian Occupational Performance Measure (COPM) Goal Sets [37] in the Exoskeleton Feasibility Study [25].

| **Child participant**<br>**(pseudonym)** | **COPM Goals** | **Functional goal area** |
| --- | --- | --- |
| Ryan<br>Outpatient | 1. Improved ability to reach for an object outside his base of support with moderate support from parents to maintain sitting. | Reach/sitting stability |
|  | 2. Improved ability to sit for 1-2 minutes without support from parents when supervised. | Sitting stability |
|  | 3. Improved independence using his walker without ankle prompts (no scissoring for 10 minutes). | Walking in own walker |
|  | 4. Improved upright posture when walking with his walker for 10 minutes. (new goal) | Walking in own walker |
| Joseph<br>School | 1. Require less support in the Rifton walker – currently needs support from PT at school to move walker | Walking in own walker |
|  | 2. Improve sitting while “floor sitting” to more than 30 seconds. | Sitting stability |
|  | 3. Would like to be able to spend longer time weightbearing on his feet. | Standing tolerance |
| Samia<br>School | 1. Walk faster in the Mustang Walker with less scissoring and less feet getting stuck (currently needs assistance with legs every 3-5 steps) | Walking in own walker |
|  | 2. Be able to load food onto a spoon with less assistance. Currently unable to load independently (usually with left hand). | Upper limb |
|  | 3. Increased time sitting independently in crossed-leg sitting (currently a few seconds). | Sitting stability |
|  | 4. Increased time to sit independently on a bench. | Sitting stability |
| Gabriela<br>School | 1. Increase sitting in cross-legged position. Currently sits for 45 seconds with hands on knees, or intermittent hands on floor. | Sitting stability |
|  | 2. In sitting, want to increase ROM of shoulder flexion to higher than 90 degrees, with increased elbow extension. | Reaching in sitting |
|  | 3. Increased targeting and increased control of elbow and waist for self-feeding using a utensil. Currently, food spills off the spoon/fork. | Feeding self |
|  | 4. Improve cruising to left and right. Would like client to be able to take a step to the side for cruising. | Cruising |
| Ana School | 1. Increase hip strength (extension) to help with dressing, i.e. putting pants on – currently knee pulls up a lot. | Hip strength (for self-care) |
|  | 2. Use more weightbearing and have less knee, hip, ankle flexion to stand. Stand with more stability. | Standing balance |
|  | 3. Improve wide steps during walking with supported walker – currently walks with scissoring. | Walking quality in own walker |
| Jasmine Outpatient | Holding up head when sitting on floor with mom. | Head control in sitting |
|  | Holding up head when supported in walker. | Head control in walker |
|  | Holding up trunk straighter when sitting on floor with mom. | Posture in sitting |
|  | Stepping with straighter legs (especially right leg). | Gait quality |
| Sophie Out-patient | 1. Improve stepping in walker. | Walking quality in own walker |
|  | 2. Improve trunk strength to allow for sitting up taller. | Sitting posture |
|  | 3. Improve ability to reach for toys when in walker. | Upper limb |
|  | 4. Improve kicking while in walker. | Function in own walker |
| Adil Out-patient | 1. Improve stepping in regular walker. Currently takes steps by pushing with both feet at a time. | Gait quality in own walker |
|  | 2. Improve use of left hand for standup assist. Currently does not use left hand for any weightbearing. | Left arm use |
|  | 3. Improve tolerance and endurance to use walker for a longer time. Currently is able to tolerate walker for 30-40 minutes. | Walking endurance in stepping walker |
|  | 4. Improve assistance for standing. Currently needs assistance to stand (50%) and will stand independently for 1 second or more. | Standing independence |
| Mark<br>Out-patient | 1. Improve ability to sit independently in cross leg position on the floor (currently needs assistance). | Sitting stability |
|  | 2. Improve ability to hold a spoon for self-feeding (holds spoon now and then throws it) | Upper limb |
|  | 3. Improve ability to steer Rifton walker with wheels unlocked. Currently can only go straight (wheels locked). | Gait quality in own walker |
|  | 4. Improve ability to stay still when caregiver is putting pants on (currently moves a lot). | Standing stability (self-care) |
| Jaden<br>School | 1. Standing at bathroom sink holding/supporting with one hand and maintain balance for a count of 3 seconds. Currently can maintain balance for 1-2 seconds so mom can wash face. | Standing balance |
|  | 2. Take 7-8 steps with one hand of mom's from bedroom toward stairs. Currently can take 5 steps. | Walking with less assistance |
|  | 3. Sit at couch with back straight while holding a book for 2 or 3 minutes. | Sitting stability |

**Table 3b.**
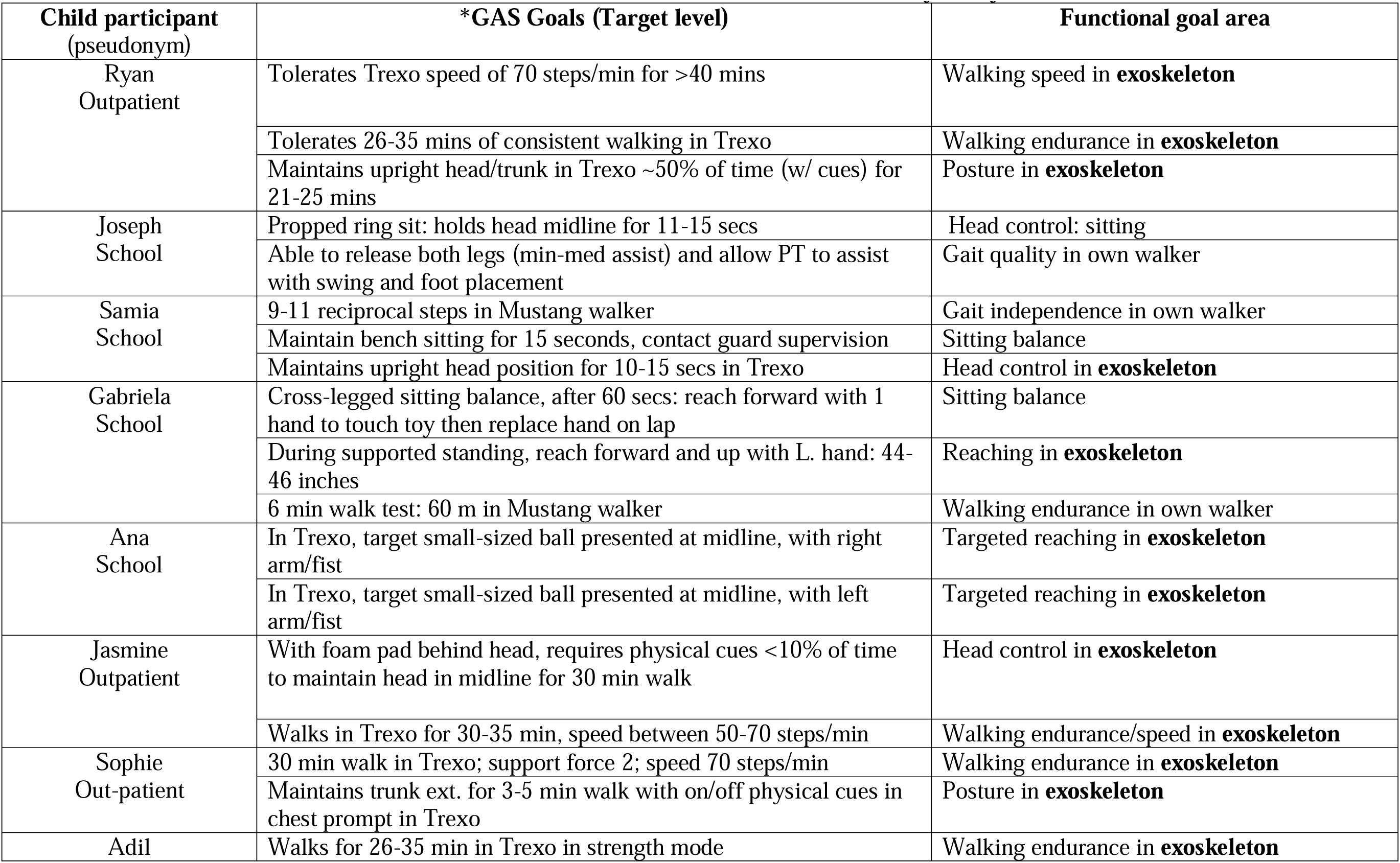
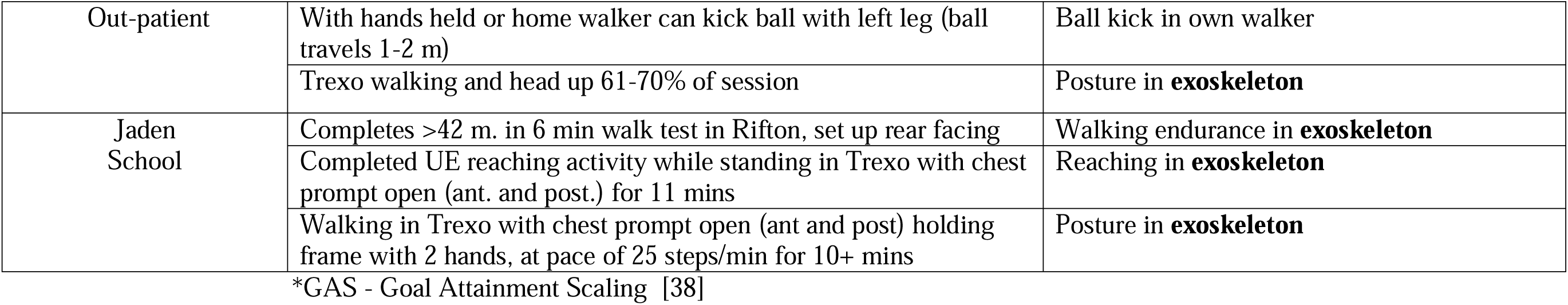
GAS Goal Sets in the Exoskeleton Feasibility Study [25].

### Study procedure

Semi-structured interviews were conducted shortly after the child completed their exoskeleton intervention block, allowing the PTs/PTAs to readily reflect on exoskeleton set-up ease, comfort, activities experienced, advantages, challenges, goals and associated outcomes, and provide recommendations toward next steps of clinical implementation (see interview guide, Figure 1). We used a paired interview strategy [39] that corresponded with our two-person teams for exoskeleton sessions where PT/PTA dyads shared child setup, activity planning and implementation, exoskeleton operation, problem solving, and child engagement facilitation. This dyadic interview approach allowed the PT/PTA pair to build on or challenge each other’s perspectives, facilitating triangulation of experiences within the interview [39].

**Figure 1.**
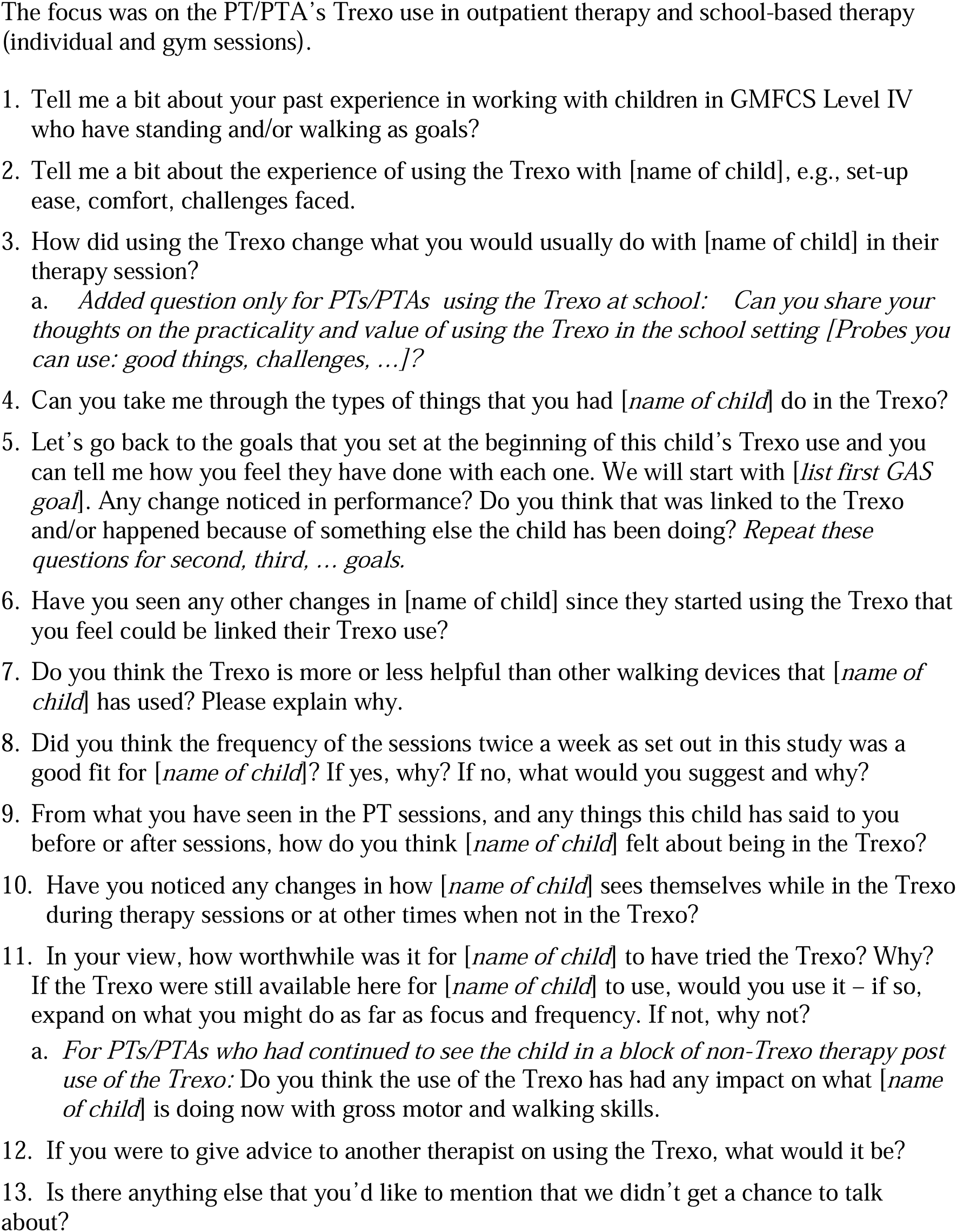
Semi-structured interview guide for PT/PTA interviewees.

Each 60-to-90-minute interview occurred over the Zoom video-conferencing platform [40]. The interviewer (CM) was a pediatric speech language pathologist with 25 years of clinical experience and no prior connection to the participants. Following CM’s interview guide orientation, a mock Zoom-based interview with senior investigator/author FVW ensured familiarity with interview questions with feedback provided on interview style and comprehensiveness.

### Positionality

Our author team was comprised of: a postdoctoral fellow (LH) with clinical experience in paediatric neurorehabilitation and research experience on robotic-assisted gait for rehabilitation of adults with spinal cord injury; a recent PhD graduate (SSB) in biomedical engineering, working on the overarching exoskeleton intervention study for her PhD dissertation; and a senior scientist (FVW) with 15 years prior experience as a pediatric physiotherapist, and lead investigator on evaluative research (quantitative and qualitative) with a gait-assisted walker (Hart Walker [41,42] and treadmill-based robotic-assisted gait device (Lokomat [43,44]) prior to their centre-based clinical introductions.

### Sample size

In the overarching feasibility study, the sample consisted of 10 child participants in GMFCS Level IV. Thus for the qualitative component, this translated into 10 treating PT/PTA dyad interviews (Table 4). There is no agreed upon sample size for qualitative studies. It has been argued that it is difficult to determine a priori [45], and post hoc decisions of adequacy need to be based on judgement of data richness (‘conceptual depth’), quality and relevance of the data (‘information power’), and strength of the data to support claims made (‘theoretical sufficiency’) [46]. As precedent, our sample size is consistent with the common range (4 to 15 interviewees) of other recent therapy-focused descriptive qualitative studies in pediatric rehabilitation [47–51].

**Table 4.** PT/PTA Interview Dyads and Corresponding Child Participants in the Exoskeleton Feasibility Study.

| **Therapist Pair (PT +PTA)** | **Therapist Interview Dyads** | **Child Exoskeleton User (pseudonym)** | **Therapy Setting** |
| --- | --- | --- | --- |
| Pair 1 | PT3/PTA1 | Ryan | Outpatient |
| Pair 2 | PT2/PTA4 | Joseph | School |
| Pair 3 | PT1/PTA2 | Samia | School |
| Pair 4 | PT2/PTA2 | Gabriela | School |
| Pair 5 | PT1/PTA4 | Ana | School |
| Pair 6 | PT4/PTA3 | Jasmine | Outpatient |
| Pair 7 | PT3/PTA3 | Sophie | Outpatient |
| Pair 8 | PT5/PTA3 | Adil | Outpatient |
| Pair 9 | PT4/PTA3 | Mark | Outpatient |
| Pair 10 | PT1/PTA4 | Jaden | School |

### Data analysis

Interviews were audio-recorded and transcribed within Zoom. Transcripts were cleaned and de-identified by a study research assistant (KT/LH) and questions referred to the interviewer (CM) for clarification. The research leads (SSB, FVW) were not involved in the interviews and had no access to the transcripts until fully de-identified.

Qualitative analysis of the interviews was data-driven using a reflexive thematic approach aligned with Braun and Clarke’s six-phase process [52,53]. Following data familiarisation through careful reading of the transcribed interviews by research student KT, interviews were coded inductively by KT using a line-by-line, phrase-based iterative approach [52]. These codes were captured within NVivo 12 software (QSR International, Cambridge, MA). Authors LH, SSB and FVW read the de-identified interviews. The initial codes were reviewed and expanded by first author LH, and coding results discussed and refined (LH, SSB, and FVW). The finalized codes were grouped into higher meaning units (categories) by LH who then built the initial themes/sub-themes [53]. Negative cases (contradictory ideas) were identified, and care was taken to include these ideas in the reporting and interpretation [54]. Thematic development occurred through a series of six team discussions (LH, SSB and FVW) through which their viewpoints and experiences informed refinements [53]. Themes were considered complete once no further explanations emerged. Dependability was supported by a member checking process [55] whereby FVW and SSB presented the themes/subthemes and quotes to the participating PTs/PTAs in a group discussion. Their ideas informed final decisions on theme wording and interpretation.

In the findings below, quotes are provided verbatim to represent the breadth of ideas expressed by the therapists. Quotes were chosen that best described these ideas and thus are not evenly distributed among participants. To maintain confidentiality, publication-specific numbers have been used for the PT/PTAs dyads (referred to below as ‘pairs’ or ‘teams’) and pseudonyms for the child exoskeleton users. Contextual clarifications added to interview quotations are shown as square bracketed text. Words with no added meaning such as “um’, “like” and “you know” have been removed to aid readability.

## RESULTS

### PT/PTA participants

Child-PT/PTA team pairings of the PTs (n=5) and PTAs (n=4) interviewed are shown in Table 4 across the sample of 10 child exoskeleton participants (five children from the out-patient program and five from the centre’s school program). Two of five out-patient children had a study PT who was their usual PT, while three children had a study PT whom they had not worked with before. All five of the school children had their usual school PT as their assigned study PT. The study’s Trexo-trained PTAs (n=4) were each assigned to a child participant(s) according to best fit with the planned exoskeleton session schedule. Characteristics of the PT/PTA interview participants are in Table 5.

**Table 5.**
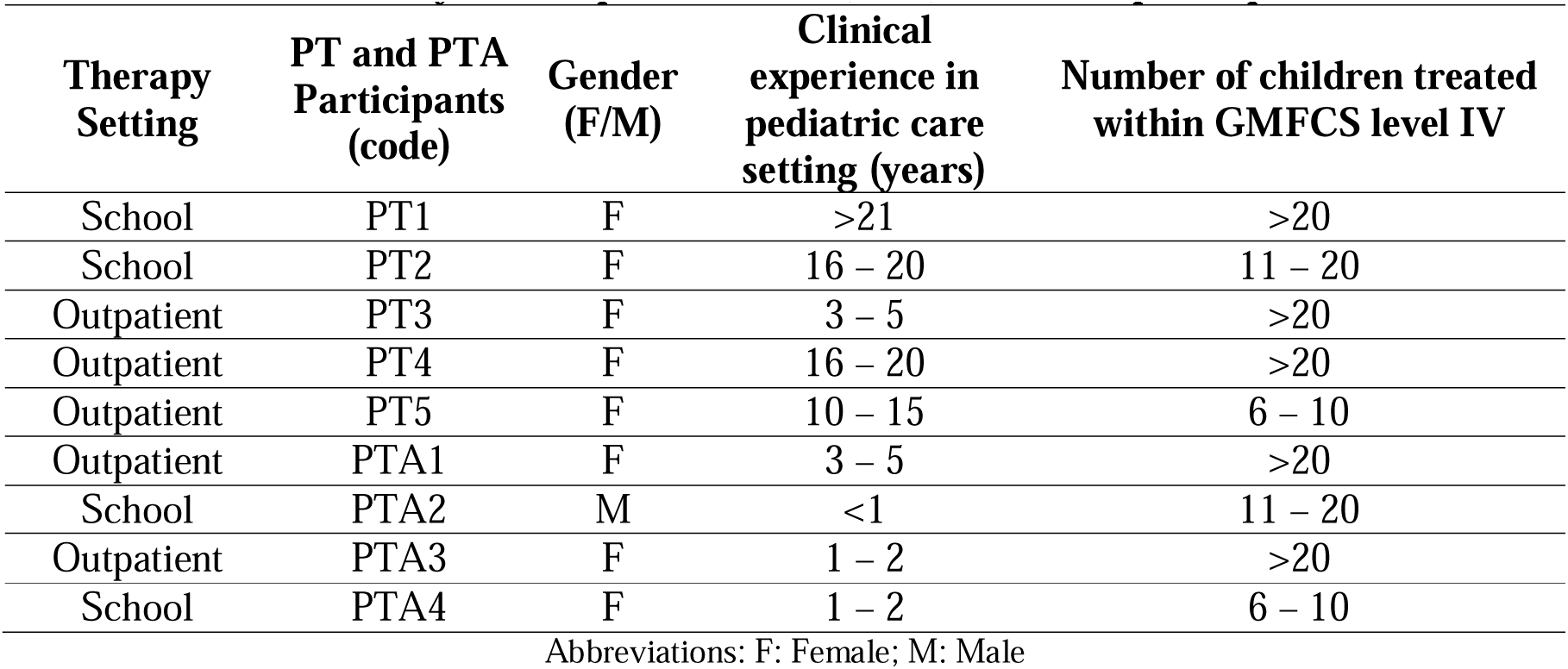
Characteristics of Physiotherapist (PT) and Physiotherapist Assistant (PTA) Interview participants.

| **Therapy Setting** | **PT and PTA Participants (code)** | **Gender (F/M)** | **Clinical experience in pediatric care setting (years)** | **Number of children treated within GMFCS level IV** |
| --- | --- | --- | --- | --- |
| School | PT1 | F | >21 | >20 |
| School | PT2 | F | 16 – 20 | 11 – 20 |
| Outpatient | PT3 | F | 3 – 5 | >20 |
| Outpatient | PT4 | F | 16 – 20 | >20 |
| Outpatient | PT5 | F | 10 – 15 | 6 – 10 |
| Outpatient | PTA1 | F | 3 – 5 | >20 |
| School | PTA2 | M | <1 | 11 – 20 |
| Outpatient | PTA3 | F | 1 – 2 | >20 |
| School | PTA4 | F | 1 – 2 | 6 – 10 |

Abbreviations: F: Female; M: Male

### Themes from the PT/PTA interviews

Three main themes were built from the interview data: 1) exoskeleton-based sessions provided additional exercise/gross motor options, and may be a valuable functionally-based adjunct to conventional therapy; 2) exoskeleton sessions were enjoyable, supporting enhanced inclusion and autonomy; 3) exoskeleton-based physiotherapy requires new clinical skills, added resources and individualized goal-based thinking. These are shown in the diagram in Figure 2 along with their related subthemes.

**Figure 2.**
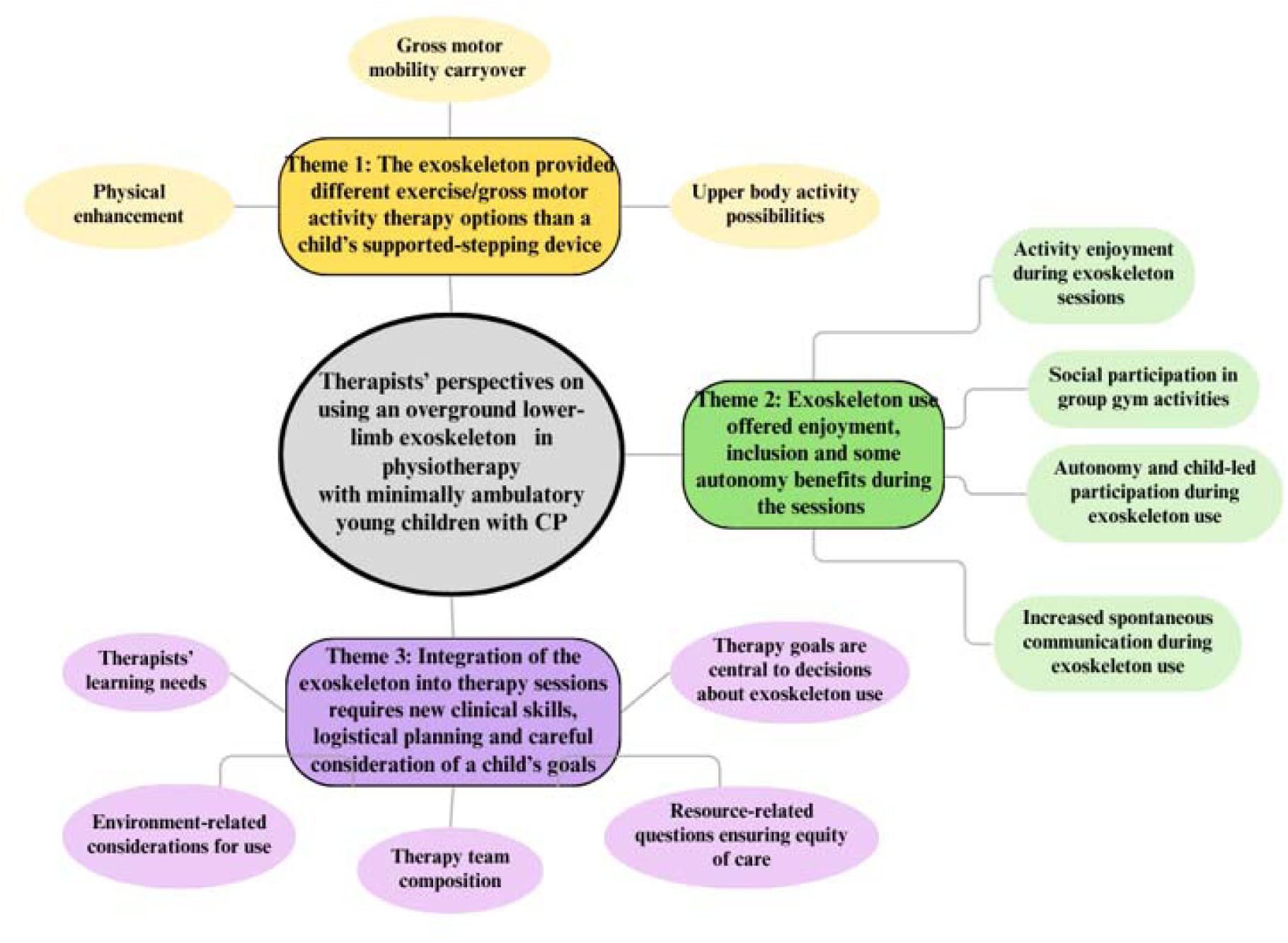
Themes and Subthemes Diagram.

#### Theme 1: The exoskeleton provided different exercise/gross motor activity therapy options than a child’s supported-stepping device

Therapists frequently described varied, and often novel exercise and functionally-based opportunities afforded by the exoskeleton, along with perceived associated outcomes, as shared below in the sub-themes. While there was a positive overall sense about these use experiences and perceived impacts, there were gross motor activity limitations and trade-offs noted compared to conventional therapy sessions, as also discussed below.

##### Physical enhancement

Therapists consistently portrayed exoskeleton-based training as an opportunity to extend beyond usual exercise and gross motor activities in the child’s supported-stepping device (e.g., Rifton/Mustang walker). There were comments from impairment viewpoint on its potential value. For example, one team noted that the reason they liked the Trexo over the Mustang “was because she does get an opportunity orthopedically [in the Trexo], a lot more extension and range of motion through her hips and knees. She’s very flexed in the Mustang.” [Pair 4: School (Sch)].

The added activity focused, walking-based exercise that a child could get was described by all teams. As one said: “The Trexo does allow her to take more steps, more symmetrical steps, and participate in a game or an activity that she wants to do on her feet. That would be very hard for her to do with her regular walker, because she just doesn’t have that consistent ability to step and the endurance also wouldn’t be there” [Pair 6: Out-patient (O-P)]. As far as what differed between use of an exoskeleton and the child’s usual stepping-support walker within a PT session with respect to session focus, one team explained:

> “If we’re going for more of an endurance type of activity, then the Trexo is gonna be more helpful for that. Maybe a strength-based activity using a walker might be better with her normal Mustang. With her Mustang she can have some control of direction herself, and I think she can get up a bit closer to some of the activities she wants to do too. But she certainly does not have the endurance to participate for as long in her own personal walker as she did in the Trexo.” [Pair 4: Sch].

Therapists described how the exoskeleton session block differed in content and focus from children’s usual physiotherapy sessions which would have been a mix of mat- and gym-based gross motor activities plus gait training in the child’s supported-stepping walker. Rather than being replacement, the exoskeleton was seen as a physical activity enhancement or adjunct to a child’s therapy program, offering a ‘more dynamic approach ‘to movement training. For one child, the team noted that within the regular therapy session:

> “There would [have been] some other things that we would work on other than just the walking component to further challenge her trunk control and her strength and her legs as well as just functional activities. So, I think [Trexo use] could be a conjunct to some of the other things that you’re working on” [Pair 6: O-P].

As well, given the set-up time to get a child into the Trexo, another team explained that exoskeleton use in a PT session needed to be entirely devoted to walking/standing activities, compared to their usual PT sessions that have been a split between mat activities and use of the child’s stepping-based walker, collectively “working on stability, transitional movements, and trying to facilitate whatever level of independence that we could.” [Pair 9: O-P].

##### Gross motor mobility carryover

Several therapists reflected on whether repeated exposure to supported stepping and upright mobility might influence motor abilities beyond the exoskeleton. Comments about this were usually made in the context of the COPM/GAS goals set for the child. While some outcomes were specific to targeted gait accomplishments in the exoskeleton, others related to new abilities in their stander, supported-stepping walker, or other devices [25].

> “Just being in that standing position [in the Trexo], he can hold his head up a little bit more and not lean so much on the makeshift headrest [when in his stander]. I think also his biking has improved. During mobility group, I noticed he’s actually able to start pushing by himself… and his head control has definitely improved on the bike.” [Pair 2: Sch].

Another team described a direct carryover to the child’s usual walker:

> “Post Trexo, we were in the gym, and we put her in her new Mustang walker, and it was a very motivating game of hide and seek, and we were all kind of taken back on the quality of the step. First, they were reciprocal. She was getting both legs going versus just the right. And they were with hip flexion, a big high step. I quickly grabbed the video, the iPad - she had a couple of instances where she took about 9 steps.” [Pair 3: Sch].

Two teams shared comments from others on potential extended benefits of children’s experiences in the exoskeleton. Specifically, one classroom teacher had commented on a child’s standing for the morning anthem, observing that: “He’ll stand up [for O Canada] from his little chair at a table with an adult, giving him a little bit of light support, but they feel his standing is better.” [Pair 10: Sch]. Another team shared that a mother reported that while less supportive than his school walker or the Trexo, her child was now using his usual walker “in the home for short distances. And he was comfortable and confident He also seemed to be able to take steps easier with hands held walking, turn his body more easily to make a turn.” [Pair 8: O-P].

##### Upper body activity possibilities

In our feasibility study, the Rifton walker frame component of the exoskeleton was positioned behind the child, allowing the child’s hands to remain free during walking and providing unobstructed access to the ‘world’ in front of them. As one pair noted: “The Trexo allows them to be hands free a little bit easier, potentially so that they can participate in activities with their family, or friends, or recreation, or in school.” [Pair 9: O-P].

Another team said that this meant that when the child was in the exoskeleton, “we did a lot of stuff with both his hands, so a lot of reaching and holding a bucket. So either grabbing snowballs (like little cotton snowballs) and putting them into the bucket, or we had a matching exercise where you match the balls in the muffin tray, which he really enjoyed.” [Pair 10: Sch]. Similarly another team observed that with this walker configuration, they could:

> “…engage much more with ‘Ana’ to actively try to hit a ball with her arms and her head was up. In the Trexo, it was interesting to be working on as much reaching and looking up and around and reaching, and midline targeting in a dynamic fashion… I wouldn’t have ever thought to do those in her Mustang which is purely for movement and activity, and for her to have that sense of independence.” [Pair 5: Sch]

#### Theme 2: Exoskeleton use offered enjoyment, inclusion and some autonomy benefits during the sessions

##### Activity enjoyment during exoskeleton sessions

Therapists frequently spoke of children’s happiness and related activity engagement during exoskeleton use, particularly in relation to being able to move comfortably through a larger environment.

> “She was quite happy, enjoyed the Trexo. I think it was the feeling of that movement and covering more floor space and moving around in gym a lot more than she would have typically been where she would have been pushed [in her Mustang].” [Pair 3: Sch] Enjoyment extended beyond walking itself, with therapists linking it to meaningful play, quality of life, and engagement. For one boy whose family who were very sport-oriented, the therapists remarked that “it was clear that it was such an amazing thing for Ryan and his mom to see him moving [in the Trexo], to be engaged, be playing soccer… In terms of quality of life, and excitement in terms of movement, that was such a big thing. And I think that’s a huge value.” [Pair 1: O-P]

This enjoyment also made a difference to children’s session tolerance and gave a chance to push to greater endurance, letting them explore new environments previously out of their reach. One therapist explained: “I think when he’s tired, he lets us know. But he is having fun. So we say, okay, we’re only gonna do 5 more minutes. He’s good with it. He’s like, okay, I can push through for another 5” [Pair 2: O-P]. Another therapist noted for a child who did more than 1000 steps in a session:

> “He fatigues quite easily within his [usual] walker. So he was able with the Trexo compared to a typical physio session, to participate more in a really fun way. He could explore different environments within the hospital, which we wouldn’t be doing [in the usual supported-stepping device]. We went to the fish tank, we went outside to the playground. It was very easy to go to different locations, which in a [usual] therapy session will probably just be in the therapy gym or one room, not really moving around too much.” [Pair 1: O-P]

##### Social participation in different group activities in school gym session

Therapists described exoskeleton use as being motivating for the child because of the expanded group interaction made possible compared to what they usually did in gym in their supported-stepping walker or when pushed in their manual wheelchair.

> “She would walk [in the Trexo] to join the class. They integrated part of it into the class where she could move between stations and participate in the activities similar to all the other kids. And there would be a relay race where they were going back and forth between activities, taking a bean bag from one side, and putting in a basket at the other side of the gym. Or obstacle courses or stationary activities where they were playing with a ball.” [Pair 4: Sch]

The exoskeleton permitted different activities in these school gym sessions than if the child was in their stepping-support walker: As one team noted, “The Trexo allowed her to use arms and legs in different activities, like playing soccer and basketball [with the class]. Where in her Mustang I feel like she was limited and she never kicked the ball ever in her Mustang because of the way she was set up in it.” [Pair 5: Sch].

One activity limitation commonly described though was with mat-based or raised surface (i.e., trampoline) gym class activities in which children could not participate, since they could not quickly be lifted out and back into the exoskeleton.

> “There are some things that he would have had to come out of the Trexo, so we either had to skip those, or if it was a preferred activity, we would promise him that at the end he could do that activity for 5 min. But then, as part of the nature of a class program where there’s multiple stations, there’s a lot of like start and stop, start and stop.” [Pair 2: Sch]

Finally, it was apparent that the different activity goals of exoskeleton use in school gym compared to individual exoskeleton sessions meant that targeted advanced planning was always needed to take full advantage of device features and possibilities. For example, the team for this same child noted that: “Overall, the gym sessions there was less of a focus on the walking and maybe more just participation and some upper extremity activities, and then the individual sessions, we really refocused on just getting as many steps in as possible for him” [Pair 2: Sch].

##### Autonomy and child-led participation during exoskeleton use

Comments in this area reflected benefits and challenges. On the one hand, therapists strongly felt they could provide meaningful choices to the child on what they would like to do in the exoskeleton session, given the wider breadth of fun options possible.

> “We would have 3 activities planned for the 40 min, and then we would ask him, ‘Would you want to do soccer [in the Trexo]?’ ‘Do you want to knock over some blocks?’ And give him the option to pick the activity he wants to do, and then try to move into the other one after 10 to 15 min. He gets time to do multiple activities. So, he leads what activities he wants to do.” [Pair 2: Sch].

Another team described greater opportunities in the exoskeleton for the child to direct activity and movement as: “She’d direct us to where she wanted to go. She could go and engage with the environment a lot more, whereas in her regular walker she isn’t really always motivated to do the walking herself, so she doesn’t get that same engagement with her classmates and the activities.” [Pair 3: Sch].

However, the underlying need for the PT/PTA to start/stop and steer the Trexo was raised as a limitation for two children who had achieved steering control in their supported-stepping walker. One PT explained that “because he can’t steer the Trexo and navigate himself, which he can do his own Rifton, you’re taking away that independence and choice making and learning of navigation. That I think may have not been as enjoyable is how much he could interact with his friends.” [Pair 10: Sch] Another PT noted gait speed limitations in one child who used the exoskeleton mainly for soccer and hallway treasure hunts in his sessions, even though he did not have fast or independent gait in his manual walker.

> “I wish that the Trexo went faster because I feel like he maxed out the speed pretty quickly that I had set that at the beginning, and then we had already maxed out 70 for most of our session. Probably halfway through our block. So I wish it went faster, because he would have enjoyed it and tolerated it.” [Pair 1: O-P]

##### Increased spontaneous communication during exoskeleton use

Several therapists described changes in children’s communication during activities in the exoskeleton, particularly with respect to vocalizations and expressive responses. This was relevant given that 7 of 10 children had restricted communication (Table 5) meaning that any momentary ‘improvement’ in their expressive communication would be important from engagement and autonomy standpoints. One clinician explained that what was very noticeable during the exoskeleton session was “when he would start babbling. I know we had one [Trexo] session where he was chatting the entire time. So I took that as a good sign. He was more engaged” [Pair 8: O-P]. Another therapist pair who had heard from a parent that the child was more vocal at home experienced the following autonomy-related verbal communication from the child.

> “I’ll never forget the day when we asked her a question during a Trexo session - I’ve known Samina 3 years, and she always just makes sounds to indicate [choices]. But I’ll never forget the morning when we asked her a question … and she responded with a ‘no’. Well, we both looked at each other, did she just say ‘no’?” [Pair 3: Sch].

#### Theme 3: Integration of the exoskeleton into therapy sessions requires new clinical skills, logistical planning and careful consideration of a child’s goals

In addition to goal-based decisions on whether and how best to embark on exoskeleton therapy, therapists all stressed that successful implementation of exoskeleton-based therapy extended beyond learning how to operate the device itself, requiring consideration of staffing models, training needs, physical environments, resource allocation, and ethical aspects of service delivery. As described below, they provided extensive comments on the training and staff resources necessary to support competent use and optimize the exoskeleton’s therapeutic possibilities.

##### Therapists’ learning needs

PTs/PTAs received structured vendor-provided training for the exoskeleton that we supplemented with goal-based/motor learning training [32,56]. However, comments about their learning needs went beyond mastering basic operation, and reflected the desire to be able to use it competently and confidently as far as setting progression and optimizing challenge and child engagement. Therapists described this competency as encompassing technical proficiency, as well as clinical decision-making on progression, engagement, safety, and goal attainment.

> “It’s a learning curve, but once you master it, you got it. As long as the fix [to a setup problem] that she [the PT] found fixed it, that’s great. If it didn’t, then we would have had to troubleshoot more. But once everything is set up for that child, it will work. You know what to do. It’s easy. You do it every single time you see the kid” [Pair 2: Sch].

Thinking about employing the exoskeleton to allow a child to progress toward their goals, one therapist explained that they “would have liked to have been a little more knowledgeable on the [Trexo’s] “endurance” versus “strength mode”. So, if I was going on to do more treatment with it, I would want to understand that and be able to change and do more of the variability in those settings.” [Pair 3: Sch]

And in line with the approach to optimize motor learning possibilities that they had been encouraged to do in this study, another therapist noted:

> “I think we learned that we could progress some of the settings on the Trexo pretty quickly. We started off the first or second session fairly conservatively. But the kids seem to adjust to the Trexo very well, and we could quickly challenge them as much as we could. [Pair 6: O-P].

##### Environment-related considerations for use

Space to navigate the Trexo was described as critical to its therapeutic use. Challenges were noted related to other people in the same area, especially in the school setting where hallways could be crowded. This meant that the therapists had to be extra vigilant in controlling the Trexo’s pace and steering.

> “During our sessions, we would be in the hallway and there’d be transitions going on. There’d be kids, adults walking in the hallway. So first, the children get very distracted and then secondly, it’s all controlled from the little tablet. So it’s kind of start [the Trexo], stop [the Trexo], look where you’re going…” [Pair 10: Sch].

Because the opportunity to take so many steps during a session was one of the observed pluses of exoskeleton use, therapists sought a diversity of environments in which to use it but ran into some obstacles to this plan. One team explained that they “did take [the Trexo] outside on a playground, but it has to still be fairly easily [accessible]. You know, door thresholds, paved area. … We did try it one time, turning it and moving it around in the elevator, and that didn’t really work all that well” [Pair 3: Sch].

Finally, exoskeleton use goes beyond what the actual sessions, and thought needs to be given as to how manage the secure space needs for the expensive device. The Trexo comes with many attachments to permit maximal adjustment to a child’s size.

> “Even just storing all the [Trexo’s] components. There’s multiple components plus storing the actual unit - where that gets kept. For this study, they kept it up in the research area, but if it was something being used exclusively in the school, I think it would be a huge storage consideration.” [Pair 2: Sch]

##### Therapy team composition

While the Trexo is designed to be operated at home/school by one adult when used purely as a mobility device, in our feasibility study, we decided on a two person (PT/PTA) team: one to steer the device from behind, and the other to engage and communicate with the child, monitor their reactions, and adjust the operating tablet’s settings to adapt to session activities. We asked the therapists for feedback on whether this two-person model should carry forward to future clinical use. They unanimously affirmed the PT/PTA team model, with discussions reflecting overarching reasons related to ease of use and therapeutic effectiveness.

> “It’s very hard to have [only] one person, because someone has to steer the whole time, and control the tablet, and then trying to get a child to engage in 40 min of activity… definitely needs more than one person.” [Pair 3: Sch]. Another therapist told us that the child’s communication needs to be considered as well and could me missed for minimally/non-verbal children in particular “if you only had one person steering and no one in front.” [Pair 9: O-P]

The need for added therapist resources was also relevant to ideas on preparing the exoskeleton for a child’s session when compared to what can be quickly done by one therapist with the child’s supported-stepping walker. Several therapists reminded us that this expensive device will need to be shared amongst children. As one explained, while supported-stepping walker is are already set up and just used by that child, “so you just put them in and then do up the straps, and then they’re good to go, [the Trexo] was a little different because there were adjustments being made to the Trexo between every use by each kid because there were multiple kids using it” [Pair 4: Sch].

##### Resource-related questions about ensuring equity of care

When asked about future integration into therapy sessions, therapists raised concerns about resource availability and feasibility of implementation and impact of equity of care, particularly in relation to the two-person care model The following ideas arose when PTs/PTAs. All spoke of concerns about allocation and availability of resources particularly if the two-person care model is applied.

> “The challenge with our [out-patient] program is resources. So if we needed two people to use the Trexo, as well as it takes a little bit longer initially for setup, or for the kids to get used to it, and then also looking at the frequency of the appointments… If we’re doing an active block of therapy with a client again for very specific goals, the most frequent we’re really seeing them is once a week. Is that enough to be in the Trexo, or do we need more than that? And if so, do we have the resources for more than that?” [Pair 6: O-P]

Very importantly, these inequity concerns extended beyond giving children fair opportunity to use the exoskeleton within their therapy, with broader implications for reduced available treatment time for other children who might not be exoskeleton use candidates but still need conventional therapy. One of the many therapists who spoke of this anticipated issue said:

> “I think it’s something we must just figure out how to use it in the school setting, while keeping interventions equitable to all the students and keeping our family’s expectations about what we would be doing with it. Which involves reviewing what that truly is from an ethical perspective. As I feel like this is the way of the future. We need to think about how we are going to equitably use something like this in a school setting. Which is going to mean more resources.” [Pair 4: Sch]

##### Goal-based considerations are paramount in deciding whether and how to include exoskeleton use within a child’s therapy sessions

As an additional link to thoughts about implementation, therapists put the exoskeleton use into a goal-based perspective that they felt needs to be at the forefront of clinicians’ thinking when planning use. As one pair noted: ““I think that it really goes back to what the family’s goals are, and how can we best work on those goals with a physiotherapist during that time, because we have limited visits and limited appointments” [Pair 8: O-P].

Therapists all indicated that decisions on when and whether to use should be shaped by the need to align use with functional goal priorities.

> “You really have to understand what your goal for that intervention is. What am I going to use this walker for? What are the advantages of using it? What do I hope it will gain for a particular goal? But I don’t think you can compare it to, apples to apples of ‘this-Walker versus that-Walker.’ … I think it [the Trexo] can be very much an adjunct to an intervention. It could be another tool in our box, of our therapy box of something that we use.” [Pair 10: Sch]

This ‘other tool in our therapy box’ perspective that was framed in the desire to address a child’s individualized goals, circles the conversation back to the Theme 1 ideas of exoskeleton use as an adjunct to usual therapy in which therapists’ overarching concern was often how to make this happen ‘within the constraints of available physiotherapy session time … while also maximizing the time of hands-on treatment in [our sessions]” [Pair 9: O-P].

## DISCUSSION

The study explored the physiotherapy team experiences, perceived benefits and challenges of overground lower-limb exoskeleton use during physiotherapy sessions with minimally ambulatory children with CP. The interviews provided direct insights into how therapists adapted to this new gait technology that was introduced in the supported context of research. Findings highlight that they considered device use through interconnected dimensions, including perceived therapeutic opportunities, child participation and engagement, alignment with child/parent/therapist goals, and practical considerations on implementation within real-world rehabilitation settings where it becomes an add-on or potential alternative to conventional physiotherapy approaches. This multidimensional perspective reflects the well-acknowledged complexity of introducing technology into rehabilitation, where clinical value must be considered alongside feasibility, accessibility, and sustainability [27,28].

While cautious about the observed outcomes they could fully attribute to the exoskeleton use, therapists showed overall enthusiasm for bringing it on board as an added physiotherapy treatment tool for primary school-aged children with CP in GMFCS IV or with similar neuromotor conditions and abilities. They also highlighted challenges related to future clinical implementation and equitable access. Each topic area is discussed below.

### The clinical role of exoskeletons may differ from other supported stepping devices, suggesting unique therapeutic applications

Therapists’ perceptions of the expanded opportunities afforded by the exoskeleton are important when considering its integration into clinical care. Children in our study had moderate to extreme difficulty propelling their supported-stepping device; however, the exoskeleton provided the possibility of higher-intensity physical activity during therapy and facilitated enjoyable walking in diverse indoor and outdoor settings. Easier and longer-duration walking has been identified as an important factor in supporting broader and more comfortable participation in daily activities [57,58]. Consistent with these findings, interview data indicated that using the exoskeleton appeared to enhance children’s motivation to engage in walking-based activities. Indeed, the frequent reference by therapists to the joy and related engagement that children showed throughout their sessions was notable. There is a strong international movement within pediatric rehabilitation to link goals and session content to the ‘F’ words for Child Development [59,60], and is now starting to become evident in other pediatric overground exoskeleton research [16]. The connection was made in our study with exoskeleton use where we repeatedly heard that the children demonstrated new <u>F</u>unctional abilities and experienced various <u>F</u>un activities in the exoskeleton while getting a chance to be physically active with peers (<u>F</u>itness and <u>F</u>riends).

Therapists felt that the exoskeleton was an enhancement to existing physiotherapy, rather than replacement, providing a new avenue to work on gait patterns/quality and functional gait-related goals that were difficult or impossible to work on through conventional approaches alone. However, limitations were experienced on what could be done in the exoskeleton. Balance, strengthening, gross motor activities related to sitting/mat/floor/balance/standing, and independent mobility training and practice (tricycle, supported-stepping/manual walker) were identified as still being essential to work on. This paralleled therapists’ comments in another pediatric study that used a powered exoskeleton walker (device not named) [29]. These observations suggest that, if thinking about introducing an exoskeleton into a child’s therapy, PTs need to take a proactive approach with families when reviewing goals and deciding the extent to which the device and focus on gait activities are suitable (or not) [61]. This process should include careful consideration of how the exoskeleton can best be integrated into the child’s broader physiotherapy plan. Delivery could be as a dedicated, functional goal-based block of exoskeleton out-patient therapy sessions (as done in our study [25] and that by Dierwechter et al. [24]), or as part of a multi-component rehabilitation program in which elements such as strengthening, balance training, and exoskeleton use might act synergistically. As our therapists indicated, and seen also in Dierwechter et al’s therapy session based study [24], potential impact seemed to go beyond walking endurance and gait quality and extend to other gross motor and functional abilities.

### Broader experiences while using the exoskeleton

A unique advantage of exoskeleton sessions was that children could use their upper limbs to handle objects and engage in play activities while walking. Unlike supported-stepping devices where child effort is directed toward self-propulsion and dynamic stability, the exoskeleton reduced these physical demands, allowing therapists to integrate mobility into meaningful functional, socially interactive activities. This aligns with contemporary task-oriented and participation-focused rehabilitation approaches which emphasize meaningful engagement within real-life contexts rather than isolated motor practice [62–64]. The arms-free advantage, identified as a critical feature in paediatric walker design for supporting child development and functional participation [4,65], is consistent with reported benefits of the hands-free, non-robotic overground Hart Walker Orthosis [41]. In that context, “standing tall” in the walker was described as a catalyst for children’s engagement and interactions within real-world activities and environments [42]. These types of interactions are not possible with stationary exoskeletons [12]. This freedom to interact with the environment while walking in the Trexo may also be advantageous from a learning perspective given the evidence of higher involvement of the sensorimotor cortex activation when children are doing activities while moving about [66].

Although not a measured outcome in our study, therapists frequently reported increased vocalization and expressive interaction during exoskeleton sessions. Upright mobility experiences may influence broader developmental engagement beyond gait practice alone [60]. These behaviours are likely attributable to increased engagement, enjoyment, social interaction, and participation within the movement-facilitated context than to improvements in communication abilities. The observations reinforce the importance of evaluating exoskeleton use via multidimensional outcomes that include communication, participation, engagement, autonomy, and overall quality of the rehabilitation experience. While not discussed by the therapists, this prompts thinking about use of a multidisciplinary approach to exoskeleton-based therapy sessions where occupational and speech therapists might also be involved.

### Device acceptability as a healthcare intervention, and considerations for implementation

As advancements in assistive technology occur, changes in service provision may follow, expanding the therapeutic toolkit available to rehabilitation professionals [67]. Acceptability (anticipated and experienced) of technology is an essential determinant of implementation success and adoption. Taken overall, this study’s findings relate to all seven facets of the Theoretical Framework of Acceptability (TFA) [68] and may help to guide next stages of implementation planning within clinical contexts. Specifically, findings discussed above reflect the TFA facets of *intervention coherence* (i.e., therapists’ growing understanding of how the exoskeleton intervention works and the fit they were able to achieve with the child’s functional gait goals), *self-efficacy* (e.g., rapidly increasing user skill for the therapists), *affective attitude* (e.g., therapists’ positivity overall about integrating the exoskeleton into their physiotherapy program given the motivation, enjoyment and engagement observed in the children), and *perceived effectiveness* (e.g., therapists’ identification of motor and broader functional areas where changes were associated with the exoskeleton). Three other TFA facets require careful consideration with respect to clinical adoption challenges, and these are: *burden*, *ethicality*, and *opportunity costs*. These are discussed in turn below.

#### i) Burden

There was, as anticipated, considerable discussion about additional time to set a child up in the exoskeleton and associated operational burden beyond that of their supported-stepping walker. This operational ‘complexity’ for overground powered exoskeletons has been identified by others as a primary barrier to adoption within out-patient settings, in which setup, technical adjustments, and therapist time were identified as important practical considerations [24,29]. Notably though, there were no comments on any reduced physical demands on the therapist during the exoskeleton sessions in comparison to the hands-on work required to facilitate a child’s gait patterns during gait training in their supported-stepping walker. The hands freeing, back-relieving advantage of fixed/mobile exoskeleton use has been described as an advantage [27]. We surmise that this might be a more notable benefit when an exoskeleton is used with older children (i.e., taller/heavier).

An exoskeleton operational drawback that was consistently mentioned was the need for therapist handling of the Trexo’s steering and start/stop/speed functions throughout the session. This is both a therapist resource burden, and could be a source of frustration for children who can independently steer a supported-stepping device and might find the Trexo experience less independent. However, in our study, there were few concerns expressed about children’s lack of directional autonomy, perhaps because therapists realized they were not independent in other aspects of mobility, i.e., none had power wheelchairs or scooters, and all required considerable help using their supported-stepping walker. As well, these children may not have been aware that they were not at least contributing to direction changes, since Trexo steering was controlled from behind (posterior handle), and direction changes were accompanied by conversations with the child about their intended destination and head/body turning cues (consistent with the motor learning approach taken),

Lastly, the distance a child can cover in this exoskeleton within a single 30 to 45 minute session (i.e., average of 1000 steps [23–25]) heightens the possibilities for broader environmental exploration. However, therapists noted that the Trexo’s size and environmental barriers in schools restricted some activities, and thus from a burden perspective, makes careful pre-planning of locations, activities, and timing essential as well as flexibility in the moment to adjust to any situations that are not navigable. Previous studies with walkers have also shown challenges of navigating less accessible environments [69–70].

#### ii) Ethicality

All therapists highlighted system-level considerations regarding equity for child (and therapist) access to exoskeleton therapy sessions if the model of service delivery used in our study (see opportunity cost section below) were to be adopted. School therapists also felt tension related to exoskeleton use in gym class, as the additional supervision required often drew their attention away from other students. These findings suggest that ethically acceptable implementation may require additional staffing resources rather than reallocating existing staff, to avoid creating inequities in service provision. These considerations parallel those expressed for use of mobile exoskeletons by physiotherapists in adult spinal cord rehabilitation, where they also felt it could be part of the intervention toolbox but needed to be integrated thoughtfully with full health system support [71], and the concept of distributive justice is well-recognized as serious issue for assistive technology overall [72].

#### iii) Opportunity costs

Cost of robotic devices goes beyond device purchase and maintenance and extends to support of staff training and clinical resource needs, and is a primary barrier to technology adoption [73,74]. Clinicians need educational materials that extend beyond safe device operation to clinical decision-making, progression, and troubleshooting [24,30,73–76]. While our therapists found the training they received was sufficient for safe use, they noted a steep learning curve and heightened cognitive workload to use, as is common with exoskeletons [24,27,68,75,76]. In our study, therapists often adopted trial-and-error and peer support approaches to adjustment of settings and progressions. As an added resource in our study, they had research team support for guidance who had direct interactions with the device manufacturer for troubleshooting, suggesting value in real world contexts to have a technology specialist and these linkages at least during early stages of implementation. Similar processes of support were described and deemed as essential by another pediatric physiotherapist group that used a Design Thinking process for Trexo introduction into their out-patient clinic [23,24], and identified as an important facilitator of adoption by Herold et al. [29] to deal with frequent set-up and technical difficulties arising during therapists’ use of a similar pediatric exoskeleton.

In our feasibility study, we provided exoskeleton sessions within our centre’s usual block treatment approach rather than ramping up session frequency over a shorter period as is often done within fixed exoskeletons to maximize neuroplasticity possibilities [18–21]. Even with this ‘usual’ block dose approach, staff availability was anticipated by the therapists to be the primary barrier to future exoskeleton integration, since all felt that a dual staff model (PT + PTA) would be required from a set-up and session activity administration/motor learning/child engagement optimization perspective. In comparison, at our centre, these primary-school-aged GMFCS Level IV children typically receive one-to-one gym-based conventional physiotherapy approach, with occasional standby assistance.

Beyond resource demands, exoskeleton purchase is a large capital investment. However, these conversations were beyond the scope of our interviews with the PTs/PTAs. These costly funding decisions demand evidence-informed comparison among different devices to justify selection of one over another [77], ideally evaluated using a cost-effectiveness framework [78].

### Study strengths and limitations

This study provides insights from PTs and PTAs on the interventional use of a lower-limb exoskeleton for young children with CP, including perceived opportunities for movement and activity participation. However, the study was conducted within a research-supported context: the research team managed device set-up and transfers between children, and PT/PTA session time was covered by the study, limiting the extent to which the resource demands of routine clinical practice were replicated [75]. The number of exoskeleton users was small and restricted to children with CP aged 4–6 years at GMFCS Level IV, which may limit applicability to other populations. Transferability to other settings should therefore be considered in light of differences in resources, funding structures, and organizational contexts, although transfer of concepts is possible [79], and the findings may inform implementation planning in similar rehabilitation settings.

## CONCLUSION

This study gave voice to the experiences and perspectives of PTs/PTAs on their first ever use of an overground exoskeleton in a school and outpatient setting with children with CP. Therapists consistently viewed the exoskeleton as a complementary intervention that could enhance, but not replace, existing physiotherapy approaches. The findings highlight the perceived value and practicalities of integrating an exoskeleton into family-centred physiotherapy practice. Future research exploring ethical considerations, models of service delivery, and clinical protocols is necessary to ensure that exoskeletons are integrated responsibly into therapy settings according to best practice principles.

## Author Approval

All authors have seen and approved the manuscript.

## Institutional Review Board Statement

The study was conducted in accordance with the Declaration of Helsinki and the Tri-Council Policy Statement in Canada. It was approved by the Research Ethics Committee of Holland Bloorview Kids Rehabilitation Hospital (no. 0523) on 14 October 2022 and the University of Toronto (no. 00044118) on 25 October 2022.

## Informed Consent Statement

Informed consent was obtained from all subjects involved in the study.

## Data Availability Statement

Data are not available on request due to privacy/ethical restrictions.

## Acknowledgments

We give special thanks to the physiotherapists and physiotherapy assistants who participated in this study. We thank Keira Tanner, undergraduate summer study, for her assistance with interview coding, and Gloria Lee, Research Manager, for her assistance with the REB and participant enrolment aspects of the study.

